# Racial and Ethnic Disparities in Prescribing of GLP-1 Receptor Agonists in the United States: A Retrospective Cohort Analysis

**DOI:** 10.1101/2024.10.28.24316312

**Authors:** Polina V. Kukhareva, Julio C. Facelli, Matthew J. O’Brien, Ram Gouripeddi, Kensaku Kawamoto, Yue Zhang, Deepika Reddy, Daniel C Malone

**Author notes:** **Corresponding author:** Polina Kukhareva, PhD, MPH, Assistant Professor, Department of Biomedical Informatics, University of Utah, Salt Lake City, UT, USA, 421 Wakara Way, Suite 108, Salt Lake City, UT 84108. **Funding Statement** This research received no specific grant from any funding agency in the public, commercial, or not-for-profit sectors. JCF and RG were partially funded by the National Center for Advancing Translational Sciences of the National Institutes of Health under Award Number UM1TR004409. MJO was partially funded by the National Institute of Diabetes and Digestive and Kidney Diseases under Award Number P30DK092949. **Contributorship Statement** PVK, JCF, KK and DCM drafted the manuscript. PVK takes responsibility for the manuscript’s content, including the data and analysis. Each author contributed substantially to the drafting or substantial revision of the paper. All authors contributed significantly to the study design, data interpretation, and manuscript writing. All authors also approved the paper for submission and agreed both to be personally accountable for the author’s own contributions and to ensure that questions related to the accuracy or integrity of any part of the work are appropriately investigated, resolved, and documented in the literature. PVK had full access to data. PVK conducted the statistical analyses.

## Abstract

**Background:** Type 2 diabetes (T2D) represents a major public health burden in the United States, with racial disparities in medication use potentially exacerbating inequities in health outcomes. This study examined racial/ethnic differences in the prescription of high-efficacy glucose-lowering medications for T2D using a large EHR network (TriNetX).

**Methods:** A retrospective cohort study included adults with uncomplicated T2D (ICD-10: E11.9), categorized as Hispanic or Latino (Hispanic) or non-Hispanic American Indian/Alaska Native (AI/AN), Asian, Black, Native Hawaiian/Pacific Islander (NH/PI), and White. Adjusted odds ratios for GLP-1 receptor agonist medications (tirzepatide, semaglutide, and dulaglutide) prescriptions in 2022-2023 were calculated by race/ethnicity, controlling for age, sex, and Charlson Comorbidity Index.

**Findings:** Among 57,320 patients included in the analysis, we observed significant racial disparities in the prescribing of GLP-1 medications. Compared to White patients, for tirzepatide, adjusted odds ratios prescriptions were 0.6 (95% CI: 0.4-0.9) for AI/AN, 0.3 (95% CI: 0.3-0.4) for Asian, 0.7 (95% CI: 0.6-0.9) for Black, 0.4 (95% CI: 0.3-0.5) for Hispanic, and 0.4 (95% CI: 0.3-0.6) for NH/PI. For semaglutide, adjusted odds ratios were 0.8 (95% CI: 0.7-0.9) for AI/AN, 0.5 (95% CI: 0.5-0.6) for Asian, 0.8 (95% CI: 0.7-0.9) for Black, 0.6 (95% CI: 0.6-0.7) for Hispanic, and 0.6 (95% CI: 0.5-0.8) for NH/PI. For dulaglutide, adjusted odds ratios were 1.2 (95% CI: 1.0-1.4) for AI/AN, 0.5 (95% CI: 0.4-0.5) for Asian, 1.0 (95% CI: 0.9-1.1) for Black, 0.9 (95% CI: 0.8-1.0) for Hispanic, and 0.5 (95% CI: 0.4-0.6) for NH/PI.

**Interpretation:** Racial disparities in high-efficacy diabetes medication prescriptions may contribute to unequal health outcomes in T2D, highlighting the need for targeted research and interventions for equitable diabetes care.

## Background

Type 2 diabetes (T2D) is a chronic, progressive disease affecting millions worldwide, with particularly high prevalence in the U.S.^1,2^ Effective T2D management hinges on pharmacotherapy, encompassing both established drugs like metformin and newer high-efficacy agents, including glucagon-like peptide-1 (GLP-1) receptor agonists.^3^ Current American Diabetes Association (ADA) guidelines recommend dulaglutide, semaglutide, and tirzepatide for individuals with T2D and no atherosclerotic cardiovascular disease (ASCVD) or chronic kidney disease (CKD) due to their superior glycemic control, weight control and cardiorenal benefits.^3^ Dulaglutide (Trulicity®), semaglutide (Ozempic®, Rybelsus®, Wegovy®), and the newest, tirzepatide (Mounjaro®), approved in 2022 as a dual GLP-1 and glucose-dependent insulinotropic polypeptide (GIP) receptor agonist, are among these recommended agents.^4^

Despite the availability of these effective medications, prescribing patterns reveal significant disparities across racial and ethnic groups, potentially contributing to unequal health outcomes.^5- 8^ Studies have shown that factors such as socioeconomic status, race/ethnicity, and geographic barriers influence access to newer T2D therapies. For instance, American Indian/Alaska Native (AI/AN), Black, and Hispanic individuals are less likely than White individuals to be prescribed sodium-glucose cotransporter-2 (SGLT2) inhibitors and GLP-1 receptor agonists.^7^ These disparities are often rooted in structural barriers, including limited healthcare access, provider biases, and gaps in insurance coverage, all of which affect equitable T2D treatment.

However, previous research has not fully captured the diversity of the U.S. population, often lacking analyses specific to AI/AN, Asian, and Native Hawaiian/Pacific Islander (NH/PI) populations, nor have they assessed the newest GLP-1 agonist, tirzepatide. This study aims to address these gaps by exploring racial and ethnic differences in the use of high-efficacy glucose-lowering medications among T2D patients in the United States, using data from the TriNetX network. By examining prescription patterns, this research seeks to clarify the scope of these disparities and provide insights to inform policy interventions that support equitable diabetes care.

## Methods

### Setting

We used the TriNetX Research Network (TriNetX, LLC. Cambridge, MA), which provides de-identified individual-level patient data and included 95 health care organizations (HCOs) across the US serving roughly 130 million patients at the time of data requisition,^9^ TriNetX has been used for various similar studies.^10^ This extensive, continuously updated database enables precise cohort construction, population analysis, and advanced statistical modeling. Data extraction for the present study was completed on June 27, 2024. This study was reviewed by the University of Utah IRB and classified as exempt from human subjects.

### Study Population

We analyzed data on adults (aged 18 years and older) with type 2 diabetes (T2D) who received glucose-lowering medications and had healthcare visits within the United States. The glucose-lowering medications analyzed included insulin, sulfonylureas, biguanides (metformin), meglitinides, thiazolidinediones, GLP-1 receptor agonists, DPP-4 inhibitors, SGLT2 inhibitors, and dual glucose-dependent insulinotropic polypeptide (GIP)/GLP-1 receptor agonists. TriNetX provided a dataset of 3,310,807 individuals from 85 healthcare organizations (HCOs). To create a more homogeneous comparison group, we restricted the dataset to individuals diagnosed with ICD-10-CM code E11.9, which designates “Type 2 diabetes mellitus without complications.” Prescription patterns were assessed only until other T2D diagnostic codes were recorded for the first time.

For longitudinal visualization, we included patients with at least one prescription for a glucose-lowering medication between 2019 and 2023. For logistic regression analyses, we focused on prescriptions from 2022-2023, as tirzepatide was approved during this period. To ensure adequate representation of minority groups, we included all individuals identifying as American Indian or Alaska Native (AI/AN) and Native Hawaiian or Other Pacific Islander (NH/PI) from the original dataset. For computational efficiency, individuals identifying as White, Hispanic, Black, and Asian were sampled. For both analyses we included patients with at least one encounter during the study period, defined as an office visit, telemedicine encounter, surgery, or lab visit.

### Covariates

Race and ethnicity were categorized into Hispanic or Latino (Hispanic) and Non-Hispanic AI/AN, Asian, Black or African American (Black), NH/PI, and White. Participants identifying with multiple racial categories were excluded because they represented the small percentage of the population. Additional covariates included age (estimated as of December 31, 2023), sex, diabetes onset (date of first recorded diabetes diagnosis), observation duration (years since first diagnosis/encounter of any type), US Census geographic region (i.e., Northeast, Midwest, South, West), last recorded HbA1c, weight, and body mass index (BMI) before December 31, 2023. The Charlson Comorbidity Index (CCI) was calculated from ICD-10 CM visit diagnosis codes preceding December 31, 2023.

### Study Measures

We performed an association study between race/ethnicity and use of new, GLP-1 glucose-lowering medications. The primary outcome was the proportion of patients receiving specific glucose-lowering medications—dulaglutide, semaglutide, and tirzepatide—during the study period. These medications were chosen because they are recommended for individuals with T2D without complications by the ADA guidelines.

### Statistical Analysis

We used mean (SD) and N (%) to describe patient characteristics. For data visualization, we used percentage of patients taking the medication of interest among patients taking any glucose-lowering medication. Logistic regression was employed to estimate the prevalence of each medication, adjusting for age, sex, and CCI. White patients were the reference group in racial comparisons.

## Results

This study included 57,320 adult patients with uncomplicated type 2 diabetes (T2D), as shown in Table 1. The cohort was predominantly female (52.7%), with a mean age of 61.92 (SD=14.08) years. Racial and ethnic composition of the cohort was predominately White patients (n=14,196), followed by Asian (n=13,982), Black (n=11,974), Hispanic (n=11,924), AI/AN (n=2,376), and NH/PI (n=2,868). White patients had the highest average age (65.5 (SD=12.98) years), whereas Hispanic and NH/PI patients were the youngest (58.8 (SD=14.52) and 58.7 (SD=13.76) years, respectively). Geographically, the US Census South region was the most represented in the sample (43.7%), with Black patients most likely to reside there (64.8%), and NH/PI patients predominantly located in the West (74.3%).

**Table 1.**
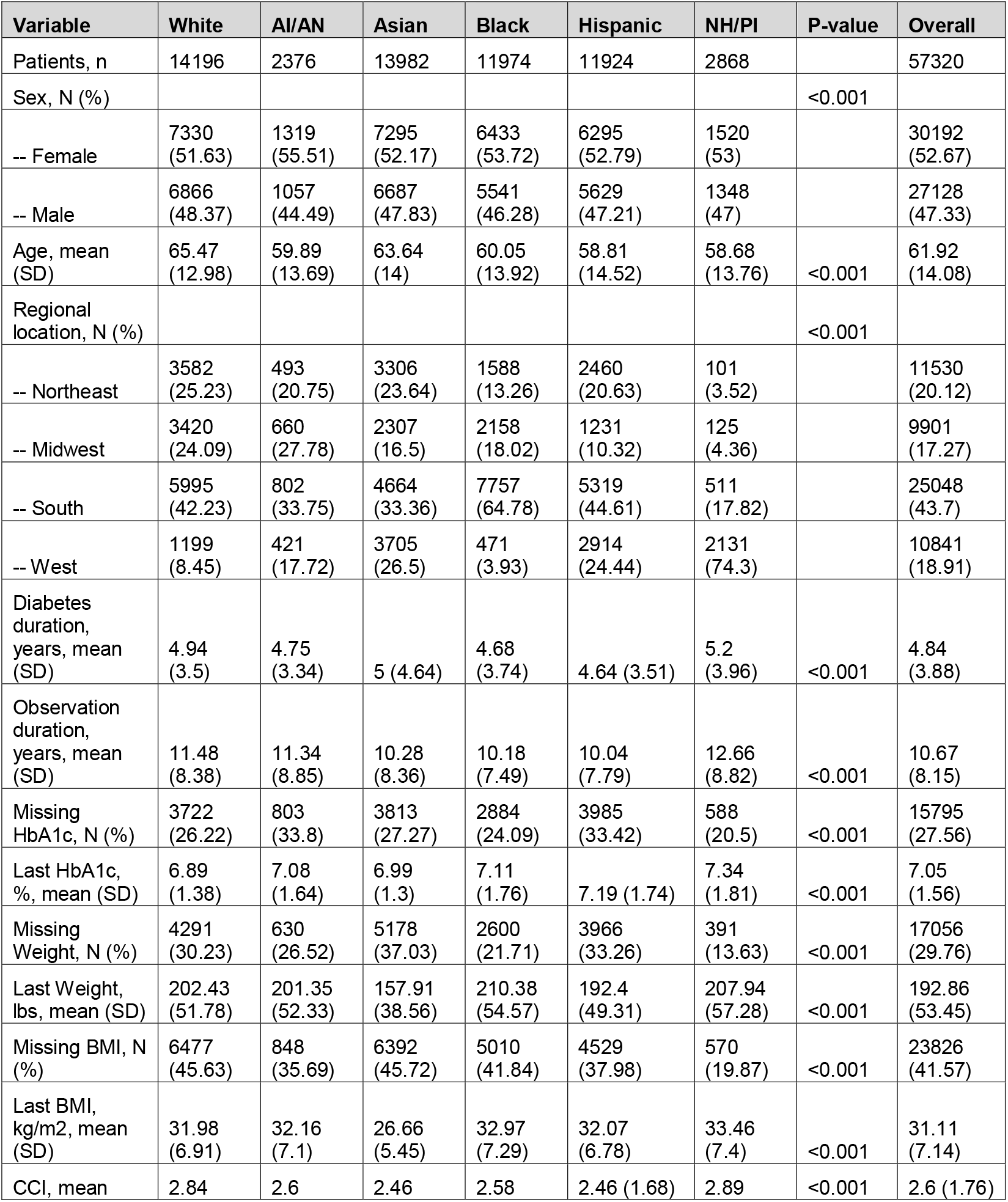
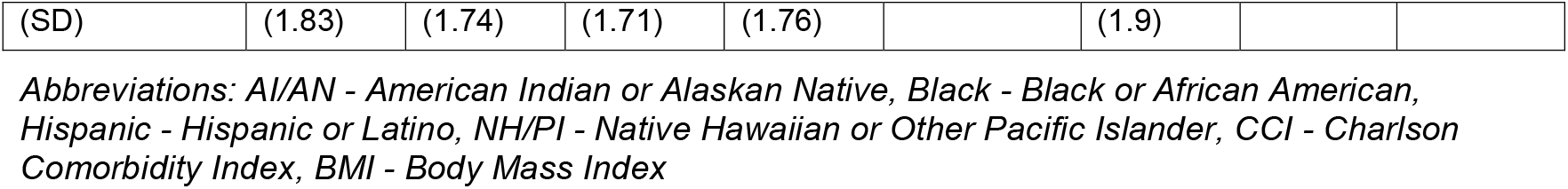
Patient Demographics and Clinical Characteristics by Racial and Ethnic Group.

| Variable | White | AI/AN | Asian | Black | Hispanic | NH/PI | P-value | Overall |
| --- | --- | --- | --- | --- | --- | --- | --- | --- |
| Patients, n | 14196 | 2376 | 13982 | 11974 | 11924 | 2868 |  | 57320 |
| Sex, N (%) |  |  |  |  |  |  | <0.001 |  |
| -- Female | 7330<br>(51.63) | 1319<br>(55.51) | 7295<br>(52.17) | 6433<br>(53.72) | 6295<br>(52.79) | 1520<br>(53) |  | 30192<br>(52.67) |
| -- Male | 6866<br>(48.37) | 1057<br>(44.49) | 6687<br>(47.83) | 5541<br>(46.28) | 5629<br>(47.21) | 1348<br>(47) |  | 27128<br>(47.33) |
| Age, mean<br>(SD) | 65.47<br>(12.98) | 59.89<br>(13.69) | 63.64<br>(14) | 60.05<br>(13.92) | 58.81<br>(14.52) | 58.68<br>(13.76) | <0.001 | 61.92<br>(14.08) |
| Regional<br>location, N (%) |  |  |  |  |  |  | <0.001 |  |
| -- Northeast | 3582<br>(25.23) | 493<br>(20.75) | 3306<br>(23.64) | 1588<br>(13.26) | 2460<br>(20.63) | 101<br>(3.52) |  | 11530<br>(20.12) |
| -- Midwest | 3420<br>(24.09) | 660<br>(27.78) | 2307<br>(16.5) | 2158<br>(18.02) | 1231<br>(10.32) | 125<br>(4.36) |  | 9901<br>(17.27) |
| -- South | 5995<br>(42.23) | 802<br>(33.75) | 4664<br>(33.36) | 7757<br>(64.78) | 5319<br>(44.61) | 511<br>(17.82) |  | 25048<br>(43.7) |
| -- West | 1199<br>(8.45) | 421<br>(17.72) | 3705<br>(26.5) | 471<br>(3.93) | 2914<br>(24.44) | 2131<br>(74.3) |  | 10841<br>(18.91) |
| Diabetes<br>duration,<br>years, mean<br>(SD) | 4.94<br>(3.5) | 4.75<br>(3.34) | 5 (4.64) | 4.68<br>(3.74) | 4.64 (3.51) | 5.2<br>(3.96) | <0.001 | 4.84<br>(3.88) |
| Observation<br>duration,<br>years, mean<br>(SD) | 11.48<br>(8.38) | 11.34<br>(8.85) | 10.28<br>(8.36) | 10.18<br>(7.49) | 10.04<br>(7.79) | 12.66<br>(8.82) | <0.001 | 10.67<br>(8.15) |
| Missing<br>HbA1c, N (%) | 3722<br>(26.22) | 803<br>(33.8) | 3813<br>(27.27) | 2884<br>(24.09) | 3985<br>(33.42) | 588<br>(20.5) | <0.001 | 15795<br>(27.56) |
| Last HbA1c,<br>%, mean (SD) | 6.89<br>(1.38) | 7.08<br>(1.64) | 6.99<br>(1.3) | 7.11<br>(1.76) | 7.19 (1.74) | 7.34<br>(1.81) | <0.001 | 7.05<br>(1.56) |
| Missing<br>Weight, N (%) | 4291<br>(30.23) | 630<br>(26.52) | 5178<br>(37.03) | 2600<br>(21.71) | 3966<br>(33.26) | 391<br>(13.63) | <0.001 | 17056<br>(29.76) |
| Last Weight,<br>lbs, mean (SD) | 202.43<br>(51.78) | 201.35<br>(52.33) | 157.91<br>(38.56) | 210.38<br>(54.57) | 192.4<br>(49.31) | 207.94<br>(57.28) | <0.001 | 192.86<br>(53.45) |
| Missing BMI, N<br>(%) | 6477<br>(45.63) | 848<br>(35.69) | 6392<br>(45.72) | 5010<br>(41.84) | 4529<br>(37.98) | 570<br>(19.87) | <0.001 | 23826<br>(41.57) |
| Last BMI,<br>kg/m2, mean<br>(SD) | 31.98<br>(6.91) | 32.16<br>(7.1) | 26.66<br>(5.45) | 32.97<br>(7.29) | 32.07<br>(6.78) | 33.46<br>(7.4) | <0.001 | 31.11<br>(7.14) |
| CCI, mean | 2.84 | 2.6 | 2.46 | 2.58 | 2.46 (1.68) | 2.89 | <0.001 | 2.6 (1.76) |
| (SD) | (1.83) | (1.74) | (1.71) | (1.76) |  | (1.9) |  |  |

Abbreviations: AI/AN - American Indian or Alaskan Native, Black - Black or African American, Hispanic - Hispanic or Latino, NH/PI - Native Hawaiian or Other Pacific Islander, CCI - Charlson Comorbidity Index, BMI - Body Mass Index

Missingness of key clinical variables is detailed in Table 1. HbA1c values were absent for 27.6% of the cohort, with the highest rates among AI/AN (33.8%) and Hispanic (33.4%) patients, while NH/PI patients had the lowest (20.5%). Weight data were missing for 29.8% of patients, with Asian patients exhibiting the highest missing rate (37.0%) and NH/PI the lowest (13.6%). BMI was unavailable for 41.6% of the population, with White and Asian patients having the highest missing rates (45.6% and 45.7%, respectively).

Figure 1 demonstrates the trend in prescriptions for dulaglutide, semaglutide, and tirzepatide from 2019 to 2023. Use of these agents increased over time with semaglutide having the most significant increase, likely due to its multiple formulations and growing evidence supporting its efficacy in diabetes and weight management. White patients had consistently higher rates of use of semaglutide and tirzepatide prescriptions than other racial groups (p=?). Although Black patients also had an increase in use of GLP-1 over time, their rates remained lower than those of White patients, with a particularly notable gap in semaglutide use in recent years.

**Figure 1.**
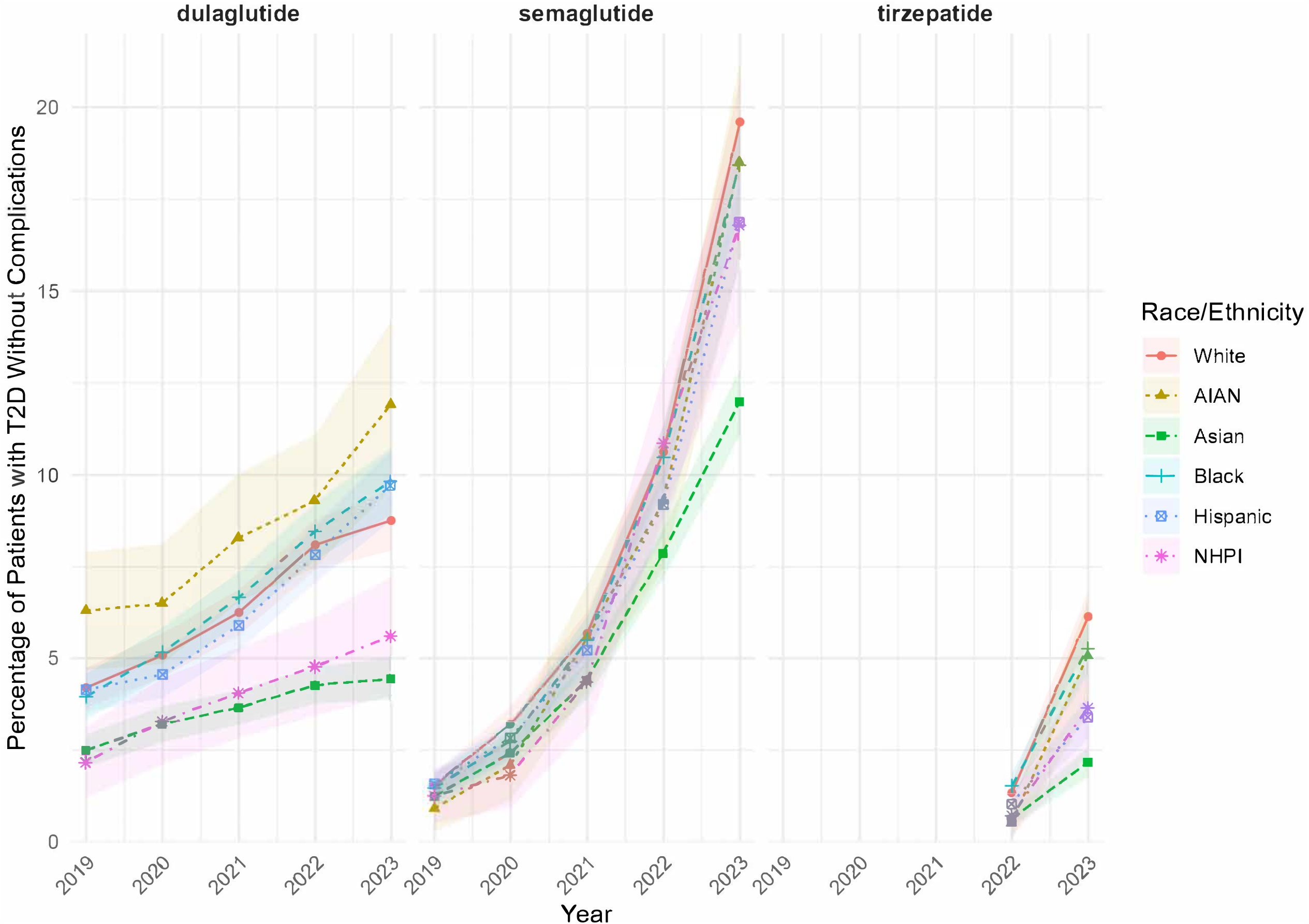
Temporal Trends in Use of Highly Effective Glucose-Lowering Medications. *Abbreviations: AI/AN - American Indian or Alaskan Native, Black - Black or African American, Hispanic - Hispanic or Latino, NH/PI - Native Hawaiian or Other Pacific Islander*

Table 2 presents logistic regression outcomes for the use of high-efficacy glucose-lowering medications. Compared to White patients, other racial groups had significantly lower odds of receiving dulaglutide, semaglutide, and tirzepatide, highlighting persistent racial disparities in access to these treatments. In 2022-2023, compared to White patients, adjusted odds ratios for tirzepatide prescriptions were 0.6 (95% CI: 0.4-0.9) AI/AN, 0.3 (95% CI: 0.3-0.4) for Asian, 0.7 (95% CI: 0.6-0.9) for Black, 0.4 (95% CI: 0.3-0.5) for Hispanic, and 0.4 (95% CI: 0.3-0.6) for NH/PI patients. For semaglutide, adjusted odds ratios were 0.8 (95% CI: 0.7-0.9) for AI/AN, 0.5 (95% CI: 0.5-0.6) for Asian, 0.8 (95% CI: 0.7-0.9) for Black, 0.6 (95% CI: 0.6-0.7) for Hispanic, and 0.6 (95% CI: 0.5-0.8) for NH/PI. For dulaglutide, adjusted odds ratios were 1.2 (95% CI: 1.0-1.4) for AI/AN, 0.5 (95% CI: 0.4-0.5) for Asian, 1.0 (95% CI: 0.9-1.1) for Black, 0.9 (95% CI: 0.8-1.0) for Hispanic, and 0.5 (95% CI: 0.4-0.6) for NH/PI.

**Table 2.** Unadjusted and Adjusted Odds Ratios for High-Efficacy Glucose-Lowering Medication Prescriptions by Racial and Ethnic Group.

| outcome | White | AI/AN | Asian | Black | Hispanic | NH/PI |
| --- | --- | --- | --- | --- | --- | --- |
| **Unadjusted** |  |  |  |  |  |  |
| Tirzepatide, % | 4.1 (3.7-4.6) | 3.2 (2.4-4.3),<br>0.8 (0.6-1.1),<br>0.11 | 1.5 (1.3-1.8),<br>0.4 (0.3-0.5),<br><0.01 | 3.8 (3.4-4.3),<br>0.9 (0.8-1.1),<br>0.33 | 2.4 (2.1-2.9),<br>0.6 (0.5-0.7),<br><0.01 | 2.1 (1.5-3.1),<br>0.5 (0.3-0.8),<br><0.01 |
| Semaglutide, % | 16.2 (15.4-17) | 15.2 (13.4-17.3), 0.9 (0.8-1.1), 0.394 | 10.6 (9.9-11.3), 0.6 (0.6-0.7), <0.01 | 15.6 (14.8-16.5), 1 (0.9-1.1), 0.369 | 14 (13.1-14.9), 0.8 (0.8-0.9), <0.01 | 13.6 (11.9-15.6), 0.8 (0.7-1), 0.018 |
| Dulaglutide, % | 9 (8.3-9.6) | 11.7 (10-13.5), 1.3 (1.1-1.6), <0.01 | 4.7 (4.3-5.2), 0.5 (0.4-0.6), <0.01 | 9.9 (9.2-10.7), 1.1 (1-1.3), 0.049 | 9.4 (8.7-10.1), 1.1 (0.9-1.2), 0.403 | 5 (4-6.3), 0.5 (0.4-0.7), <0.01 |
| **Adjusted** |  |  |  |  |  |  |
| Tirzepatide, % | 3.8 (3.4-4.3) | 2.4 (1.8-3.3), 0.6 (0.4-0.9), <0.01 | 1.2 (1-1.5), 0.3 (0.3-0.4), <0.01 | 2.8 (2.4-3.2), 0.7 (0.6-0.9), <0.01 | 1.6 (1.4-1.9), 0.4 (0.3-0.5), <0.01 | 1.5 (1.1-2.2), 0.4 (0.3-0.6), <0.01 |
| Semaglutide, % | 16.6 (15.8-17.5) | 13.3 (11.6-15.2), 0.8 (0.7-0.9), <0.01 | 9.8 (9.2-10.5), 0.5 (0.5-0.6), <0.01 | 13.4 (12.6-14.3), 0.8 (0.7-0.9), <0.01 | 11.1 (10.4-11.9), 0.6 (0.6-0.7), <0.01 | 11.4 (9.9-13.1), 0.6 (0.5-0.8), <0.01 |
| Dulaglutide, % | 9.3 (8.7-10) | 10.9 (9.4-12.7), 1.2 (1-1.4), 0.070 | 4.6 (4.2-5.1), 0.5 (0.4-0.5), <0.01 | 9.2 (8.5-9.9), 1 (0.9-1.1), 0.733 | 8.3 (7.6-9), 0.9 (0.8-1), 0.029 | 4.6 (3.6-5.8), 0.5 (0.4-0.6), <0.01 |

Values for White are expressed as estimated mean with 95% confidence intervals (CIs). Values for AI/AN, Asian, Black, Hispanic, and NW/PI are reported as estimated mean with 95% CI, followed by odds ratios (ORs) with 95% confidence intervals (CI). All values were calculated using logistic regression. "Adjusted" values are adjusted for age and sex.
Abbreviations: AI/AN - American Indian or Alaskan Native, Black - Black or African American, Hispanic - Hispanic or Latino, NH/PI - Native Hawaiian or Other Pacific Islander

## Discussion

In this analysis, individuals identifying as White were more likely to receive prescriptions for GLP-1 receptor agonists than individuals from non-White racial and ethnic groups. Individuals in all non-White groups had lower odds of receiving tirzepatide and semaglutide prescriptions, while no significant differences were observed in dulaglutide prescriptions for individuals identifying as AI/AN, Black, or Hispanic compared to White individuals. However, those identifying as Asian and NH/PI were significantly less likely to receive dulaglutide than White individuals. These findings underscore persistent racial and ethnic disparities in access to newer, high-efficacy diabetes medications, which may contribute to differences in treatment outcomes across these groups.

Notably, individuals identifying as Asian had especially low odds of GLP-1 receptor agonist prescriptions. Asian Americans have a higher prevalence of type 2 diabetes than the general population and are at increased risk for diabetes at lower BMIs.^11^ Moreover, this population is more likely to accumulate fat in visceral adipose tissue, which is metabolically harmful and a significant risk factor for diabetes. However, the lower prescription rates for GLP-1 medications, including tirzepatide and semaglutide, among Asian individuals may stem from clinical perceptions influenced by BMI. Providers may underestimate diabetes risk in this group if they are guided primarily by BMI cutoffs, overlooking the unique risk factors that characterize diabetes development in Asian populations.

The clinical implications of these disparities are substantial. Limited access to optimal diabetes pharmacotherapy is associated with poorer glycemic control and increased risk of severe complications, including cardiovascular disease, chronic kidney disease (CKD), and premature mortality.^8,12^ Previous studies have documented that Black and Hispanic populations bear a disproportionate burden of cardiovascular mortality and CKD.^8,12^ Our findings align with these reports, suggesting that disparities in diabetes management may further exacerbate these outcomes by restricting access to treatments capable of reducing complications and improving overall health.

These results are consistent with broader healthcare disparities documented in the United States, where structural barriers—including socioeconomic status, insurance access, and geographic location—limit access to advanced treatments among minority populations. Provider biases and variations in healthcare delivery across institutions add to these disparities, mirroring patterns seen in cancer treatment, cardiovascular care, and maternal health. Addressing these inequities necessitates a multifaceted strategy, including policy measures to improve insurance coverage, initiatives to promote equitable prescribing practices, and community outreach programs to enhance medication access in underserved populations.

This study provides unique insights into diabetes pharmacotherapy trends among populations frequently underrepresented in research, including Asian, AI/AN, and NH/PI individuals. By leveraging the TriNetX database, which aggregates clinical data from a diverse network of healthcare organizations, this study captures a broad sample that includes substantial representation from these groups. This robust dataset allows for detailed subgroup analyses that are often unattainable in smaller or less diverse studies. While prior research has addressed racial disparities in diabetes treatment, smaller groups like AI/AN and NH/PI individuals are often excluded due to sample size limitations. This study, by contrast, enables an assessment of treatment patterns within these populations, illuminating their specific healthcare needs and underscoring disparities in access to high-efficacy medications. The separate analysis of Asian patients allows for the capture of critical nuances in diabetes risk profiles and treatment responses unique to this diverse population, informing more precise, equitable diabetes care.^11^

### Limitations

Using EHR data from sources like the TriNetX network provides valuable insights into patient care and treatment patterns but is not without limitations. First, EHR data may not fully represent the broader population, as some demographics may be underrepresented based on the locations and types of healthcare facilities contributing data to the network. This could limit the generalizability of findings, especially if specific racial or ethnic groups are disproportionately represented or excluded. Second, EHRs often has documentation gaps, including missing lab results, medication details, or demographic information, which may impact the reliability of the findings.^13,14^ Variability in data entry and coding practices across providers and institutions can introduce inconsistencies, potentially leading to bias or misclassification of diagnoses and prescriptions. Additionally, we did not have access to certain covariates, such as insurance status, which could further inform understanding of prescribing behaviors.

## Conclusion

This study identified significant racial and ethnic disparities in the prescription of high-efficacy glucose-lowering medications, specifically dulaglutide, semaglutide, and tirzepatide, with non-White patients consistently exhibiting lower odds of receiving these treatments compared to White patients. Such inequities in access to diabetes pharmacotherapy are of particular concern as they potentially contribute to poorer glycemic control and heighten the risk of serious complications, including cardiovascular disease, CKD, and premature mortality. These findings underscore the pressing need for targeted interventions to address these disparities, ensuring equitable access to effective diabetes treatments across all racial and ethnic groups.

## Data Availability Statement

The data underlying this article cannot be shared publicly due to TriNetX policies.

## Acknowledgements

During the course of preparing this work, the author(s) used **ChatGPT 4.0** for the purpose of **text editing**. Following the use of this tool/service, the author(s) formally reviewed the content for its accuracy and edited it as necessary. The author(s) take full responsibility for all the content of this publication.

